# Instrumented dual-task tests for concussion assessment in Ice Hockey

**DOI:** 10.1101/2024.10.25.24315523

**Authors:** Frederic Meyer, Nicolas Baehler, Dario Sciacca, Lea Chabrowski, Mathieu Falbriard, Anisoara Ionescu

## Abstract

**Background:** Sport-related concussions are a public health concern and can lead to long-term health impairment. Nevertheless, assessing concussions in spots such as ice hockey can be challenging during games. Dual-task (DT) tests, quantifying interference between cognitive and motor performance, have been used to detect cognitive impairment after a concussion.

**Methods:** in this work, 114 Swiss elite ice hockey players performed DT tests at regular interval during the season and within the days following a diagnosed concussion. The DT test consisted of a static balance and a self-paced walking, both combined with counting backward by 3. In total, 265 DT tests were performed, including 38 tests between 1 day and 2 weeks after the 15 diagnosed concussions.

**Results:** Cognitive abilities during gait were mainly affected after a concussion. A DT performance metric was defined based on features significantly associated with worsening DT performance after the concussion (*p*< 0.001, effect size *d* = 1.50). Finally, an individual and a global model were proposed to estimate the risk of concussion based on a DT test outcome.

**Conclusion:** These tests provide objective insights to the medical staff, ensuring a secure and trusted decision-making process when deciding if the player should be removed from the game for recovery. Dual-task tests could also improve the return to play evaluation by detecting remaining cognitive deficiencies even in the absence of symptoms.

## 1. Introduction

Mild Traumatic Brain Injuries (mTBI) often interchangeably called concussions, are one of the prevailing injury risks in contact sports such as American football, ice hockey and rugby ^1–3^. In ice hockey, the incidence of mTBI is estimated to be between 5.8 and 6.1 per 100 games ^4^.

Sport-related concussions (SRC) are caused by a direct blow to the head, neck or body, resulting in a sudden linear or rotational acceleration of the head. Such an event causes an excitation of the neurotransmitters in the brain that induces a sudden metabolic demand and can lead to axonal injury, blood flow and brain inflammation ^5^. The resulting symptoms vary among individuals and can appear instantaneously or up to a few hours after the incident. The type of symptoms depends on several factors such as the location and severity of the impact, as well as the age or sex of the patient ^5^. Common symptoms include concentration problems (in 40-90% cases), headaches (in 70-80% cases), vertigo, dizziness and balance issues (in 37-81% cases), neck pain (in 20-50% cases), visual problems (in 20% cases), fatigue (in 20-50% cases) and autonomic dysfunctions (in 20-60% cases) ^6^.

The majority of the assessment methods used today are based on a questionnaire-type diagnosing, which covers and evaluates different aspects of the condition. The Sport Concussion Assessment Tool 6 (SCAT6) ^7^, Concussion Recognition Tool (CRT5) ^8^, Standardized Assessment of Concussion (SAC) ^9^, and Balance Error Scoring System (Modified BESS) ^10^ are a few standard assessment tools. SCAT6 requires more than 15 minutes to be conducted and requires medically trained personnel to evaluate and score the questionnaire.

Once a diagnosis is made, the medical staff sets up a return-to-play procedure (RTP). The International Ice Hockey Federation (IIHF) published a dedicated protocol in six steps ^11^. After a mandatory 24-hour to 48-hour rest period, each step should be performed with a minimum of 24 hours in between. If a step is not validated, it should be repeated the day after. The whole procedure takes, therefore, a minimum of one week, and the player is cleared for RTP when all the clinical symptoms have disappeared. Similar RTP protocols are applied in other sports, but recent work showed limitations to this process, as it has been observed that players who sustained a concussion were more prone to suffer a second musculoskeletal injury, suggesting that proper recovery is oftentimes not achieved with symptoms disappearance ^12^. Recently, a literature review highlighting the absence of neuro-cognitive testing in the RTP protocols hypothesized that focusing only on clinical symptoms could lead to too early RTP clearing, as athletes could still suffer from neurocognitive impairment ^13^. Moreover, it was recently observed that athletes with lower neurocognitive functions had more anterior cruciate ligament injuries ^12^. Testing neurocognitive functions before the RTP seems to be an essential step reducing the risk of future injuries on athletes.

Dual-task (DT) tests have been used to detect concussions in contact sports ^14^. The test consists of performing a cognitive task while concurrently performing a motor task. To complete both cognitive and motor functions simultaneously, attention needs to be shared, leading to increased attentional demands and inducing interference between the two tasks ^15,16^.This interference, called cognitive-motor or DT interference, reduces performance in one or both tasks ^18,19^. The specific choice of attentional resource allocation is called prioritization. This phenomenon emerges spontaneously when no explicit instruction is given to prioritize one task over the other. Healthy adults tend to naturally adopt a "motor first" approach when subjected to a DT gait situation ^20^.

Cognitive tasks such as counting backwards, list of words recall, reaction time, auditory Stroop test and verbal fluency task are commonly used in DT assessment ^17^. As motor tasks, dynamic balance during walking or keeping static postural balance is often used ^16,17^. Static balance is most commonly evaluated using force plates, but several studies have also used inertial measurement units (IMU) ^21^ and even smartphone IMUs ^22,23^. While IMUs can be placed on different body parts, they are usually placed in the lower lumbar region of the participant, which is considered to track the subject’s center of mass optimally ^24^. Gait spatio-temporal parameters can also be determined using IMUs to quantify gait quality. Walking speed and variability were affected up to ten days after a concussion before returning to normal ^25^.

Therefore, this study aims to validate a DT assessment procedure based on the use of smartphone-embedded sensors to record raw DT data and employ data processing algorithms to extract metrics for a direct and quick evaluation of ice hockey players.

## 2. Methods

### 2.1 Participants

In total, 114 male professional ice hockey players (age: 27.6, ± 4.6 years, height: 182.5 ± 6.1 cm, weight: 85.8 ± 7.5 Kg) active in both the highest and the second-highest Swiss ice hockey leagues participated in the study. They were informed of the benefits and risks of this investigation before giving their written informed consent. Our Institutional Ethics Committee previously approved the experiment (CCER-VD 2023-00039) and complied with the Declaration of Helsinki.

### 2.2 Experimental setup

The protocol makes use of one cognitive and two motor tasks. The cognitive task consists of continuously subtracting 3 from a randomly generated number between 300 and 900. The first motor task consists of keeping balance in a bipedal stance, looking at a sticker on a wall at 1.6 m height and 2.5 m away. The second motor task consists of self-paced walking around two marks placed at a 10 m distance. This result in five different tests: (1) The single-task (ST) cognition test: 60 seconds (s) in a sitting position and counting backwards in steps of three. (2) The ST balance test: 60 s in bipedal stance. (3) The ST gait test: 60 s self-paced walking with 5 s still phase at the end. (4) The DT balance: standing up in bipedal stance and counting backwards in steps of three for 60 s. (5) The DT gait: self-paced walking for 60 s while counting backwards in steps of 3.

Regarding the cognitive task, serial subtractions by 3 is a practical cognitive task in the context of DT testing ^17^. It belongs to a category of cognitive tasks often referred to as question and answer-type tasks, which are effective for DT testing ^26–28^. Moreover, it is convenient as it does not require any additional gear, and does not require inputs from the instructor during the 60 seconds. Picking different starting numbers is also very versatile, and the participants cannot develop disruptive strategies to improve their performance.

### 2.3 Equipment

Two smartphones (iPhone 8, Apple, USA), two wireless microphones (Callstel HZ-2789, Pearl, Switzerland) and a laptop (T14s, Lenovo, China) were used for this experiment. The first smartphone was placed in a waist bag and strapped to the participants’ lower back, in the L5 region. The height of the smartphone’s centre above ground was measured before starting the test. The Sensor Logger app (Kelvin Choi, Sensor Logger, App Store, USA) was used to record the smartphone’s 3D accelerometer (100Hz) and 3D gyroscope (100Hz). The audio was also recorded using the wireless microphone placed on the participant’s collar. To facilitate the data collection process, we used a custom terminal application (Bearmind, Switzerland) to guide the instructor through the different stages of the test. The application aims to store all the necessary information about the trial, use timers to ensure the correct durations for each task and record the timestamps at the beginning and the end of each test. The system also recorded a backup audio file using the second wireless microphone in case of smartphone audio recording failure.

### 2.4 Data processing

#### 2.4.1 Cognitive task

The audio of the cognitive test phase and the two DT test phases was first extracted. The experimenter then transcribed each file to determine the number of correct answers *(correct number)*, the rate of correct answers *(correct rate)*, the mean and variability of the answers duration (*durations mean and std*), and the mean and variability of the pauses between answers (*pause mean and std*). The different phases of the test were also determined using the audio file.

#### 2.4.2 Gait task

A Principal Component Analysis (PCA) was performed on the acceleration data of the first two seconds of the walking period to determine the smartphone’s orientation. The first component of the PCA indicates the vertical axis, as most of the acceleration is due to gravity. The second component is attributed to the antero-posterior (AP) axis, as the participant starts his motion in the AP direction. The third component is finally attributed to the medio-lateral (ML) axis, where lower accelerations are visible ^29^.

Once phases and sensor orientation were determined, walking turns were detected using a 10 degree-per-second threshold on the angular velocity norm. Different gait metrics were then computed on each of the straight walking bouts. *Cadence* was calculated based on the detection of step initial contacts and provided for a granularity of a one-second window. A Hampel filter was applied to the step frequency to remove outliers ^29^. The *step length* was computed using the method proposed by Zijlstra *et al.* ^30^, using an inverted pendulum to model human gait:

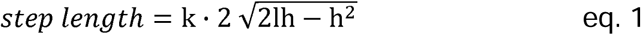

Where *k* is a correction factor (*k =* 1.25), *l* is the pendulum length (i.e. the sensor height), and *h* is the vertical displacement of the center of mass obtained by double integration of the vertical acceleration component during step duration. The *step length* estimation is also computed at a granularity of one second windows. The gait *speed* is finally calculated as half the *step length* times the *cadence* per second.

#### 2.4.3 Static Balance task

A set of metrics were computed on both medio-lateral (*ML*) and antero-posterior (*AP*) acceleration components to assess the static balance performance. We choose these metrics for their relevance in characterizing body sway. They are indeed commonly used to assess postural sway in patients with neurological disorders ^21^, making our research directly relevant to real-world scenarios (Table 1). The frequency below which 95% of the power spectra of the acceleration signal is present (*F95)* and the *spectral centroid* belong to the frequency domain features, whereas all the other metrics belong to the time domain features ^31^.

**Table 1:**
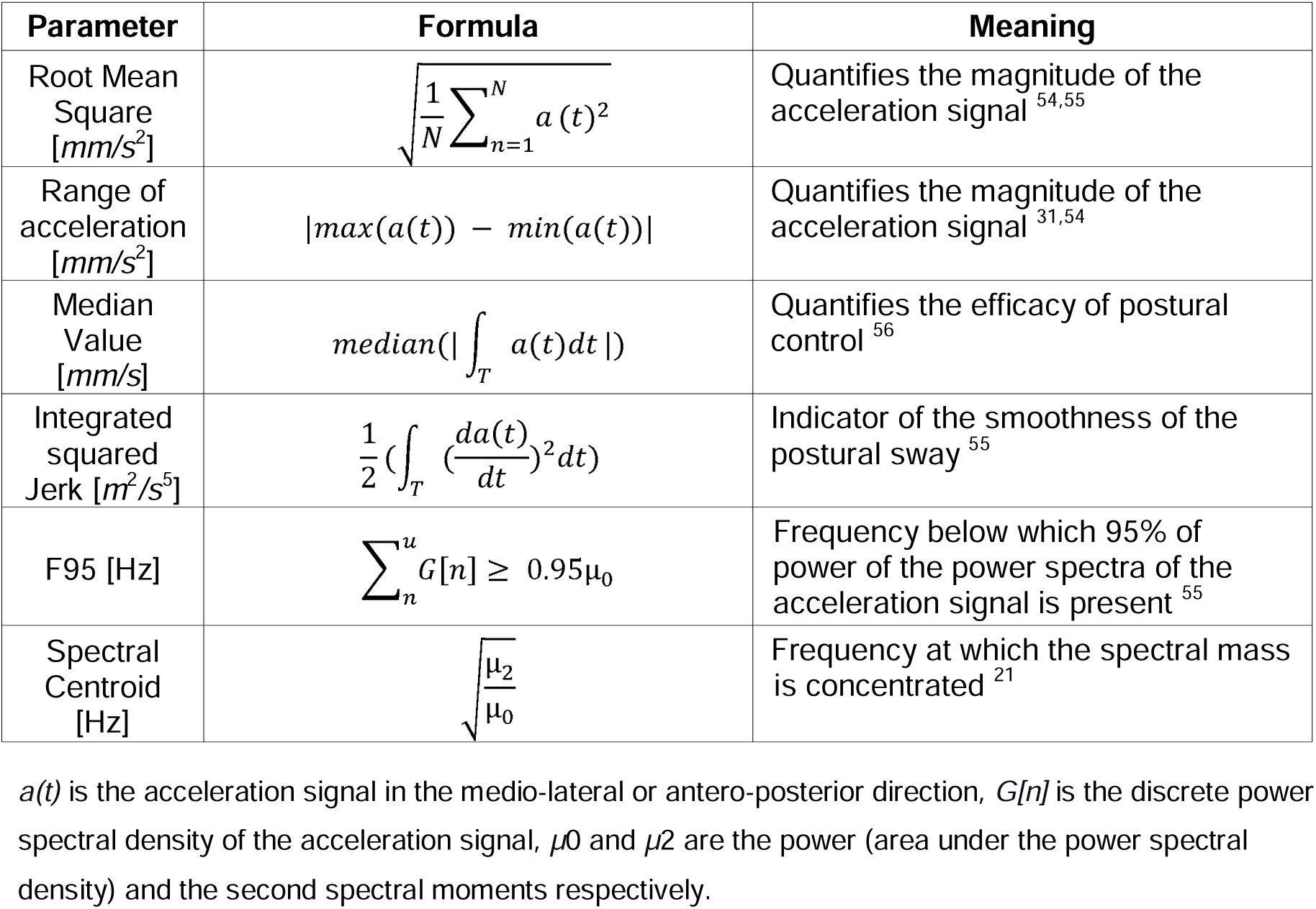
Body sway metrics computed from the acceleration signals recorded with the Smartphone IMU.

The acceleration signals measured with the Smartphone were filtered with a 4^th^-order Butterworth lowpass filter with a cutoff frequency of 3.5 Hz to compute the linear metrics ^31^. This lowpass filtering allowed the removal of tremors, if any. Before computing the *acceleration rang*e with the formula shown in Table 1, we removed the outliers by only taking the values that were between 1% and 99% of the acceleration distribution.

In order to compute the *median value* and the *integrated squared jerk* with the formulas shown in Table 1, the acceleration signals were filtered with a 4^th^-order Butterworth high pass filter with a cutoff frequency of 0.5 Hz before integration to attenuate the drift effect^31^. The *spectral centroid* was computed based on a function from the Matlab Audio Analysis Library ^32^. It provides a noise-robust estimate of how the dominant frequency of a signal changes over time.

#### 2.4.4 Dual-task effect

To quantify the cognitive-motor and the cognitive-balance interferences under DT conditions, the DT effect has usually been computed as the difference between the DT and the ST performance divided by the ST performance ^33^. In the current work, we also normalized the DT effect using the Bland-Altman method (eq. 2) ^34^. This process removes extreme values when the ST performance is close to zero.

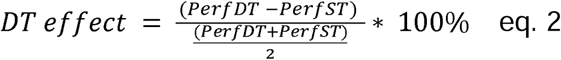

Where *PerfDT* is the performance under DT conditions, and *PerfST* is the performance under ST conditions. For most features, a smaller value indicates a better performance (e.g. the variability, the *RMS* and other metrics related to the balance). The corresponding effect is, therefore, positive when DT performance has higher values. For the features where a higher value represents better performance (i.e. *correct number*, *correct rate*, *speed* and *step length*), the sign was inverted to keep positive effects when the performance under DT condition is worse than in ST. Therefore, positive values of the DT effect always indicate better performance in ST than in DT ^35^.

### 2.5 Analysis

#### 2.5.1 Single-task vs dual-task

The difference between the ST and the DT was assessed using the paired Wilcoxon signed rank tests applied to each feature, as normal distribution was not observed for each feature. A significant difference was set for *p*< 0.05.

#### 2.5.2 Intra-subject variability

Players performed multiple DT tests during the season. In order to highlight a possible learning effect of the test, the variability between the first and the second test was compared if the tests were performed in the first month of the season. Tests conducted later in the season were not considered in the analysis to remove possible fatigue effects that can happen during the season. The pre-season baselines were compared with those performed at the end of the season (more than 18 weeks after the beginning) to highlight possible long-term fatigue effects. Paired Wilcoxon signed rank tests were applied on each feature effect with a significant difference set for *p* < 0.05. Cohen’s effects size *d* were also calculated ^36^. Values from less than 0.2 indicate a very weak effect, from 0.2 to 0.5: weak, 0.5 to 0.8: moderate, 0.8 to 1.2: strong, 1.2 to 2: very strong and more than 2: extremely strong ^37^.

#### 2.5.3 Intra-subject concussion analysis

For players that sustained a concussion assessed by the club medical doctor during the season, paired Wilcoxon signed rank tests were applied on each feature effect to compare their baseline test with those performed right after the concussion. Moreover, the athlete’s baseline was also compared to a DT test performed five to seven days after the concussion to see if the players were still affected. A significant difference was set for *p* < 0.05. Cohen’s effects size *d* was also calculated. Features with statistically significant differences were added and averaged to determine the total DT effect. This total DT effect was then subtracted to 100 to calculate the DT performance index (in %). A performance lower than 100% indicates an overall DT cost. The normal distribution of the baselines and the concussion groups was assessed to build a specific concussion risk model. If normal distribution of both groups is observed, the probability density function (*PDF*) of the standard normal distribution of the DT performance index will be used (eq.3) ^38^. Otherwise, the empirical cumulative distribution function will be used to build the concussion risk model.

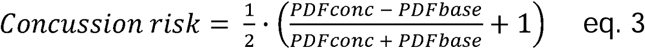

#### 2.5.4 Inter-subject concussion analysis

In actual-life situations, some athletes will sustain a concussion during the season but will not have performed a baseline before the season starts. It is, therefore, interesting to compare the group that performed a DT test right after sustaining a concussion with the group of players that performed a baseline pre-season but did not sustain a concussion. Therefore, the total DT effect of the two groups was compared with a Wilcoxon signed rank test and a significant difference set for *p* < 0.05. Cohen’s effect size *d* was also calculated. A global model to estimate concussion risk was also built with the distribution of the pre-season baselines (without the baselines corresponding to athletes that sustained a concussion later in the season), as described in eq.3.

Finally, the concussion group was compared to the group of athletes who did not sustain a concussion during the season and made a baseline at the end of the season. Paired Wilcoxon signed rank tests were applied on each feature effect to compare the baseline test with the test performed right after the concussion. A significant difference was set for *p* < 0.05. Cohen’s effects size *d* were also calculated.

## 3. Results

In total, 265 DT tests were performed, including 38 tests between 1 day and 2 weeks after the 15 diagnosed concussions.

### 3.1 Single-task vs dual-task

For the gait features, significant differences between ST and DT performance were observed for the *cadence mean, step length mean, turning time mean and speed mean*, estimated over the walking path (all *p* < 0.05). The features concerning the variability of the different gait parameters were not significant (all *p* > 0.2). For the cognitive features, only the *correct rate* showed a significant difference between ST and DT during gait (*p* = 0.014) (Figure 1).

**Figure 1:**
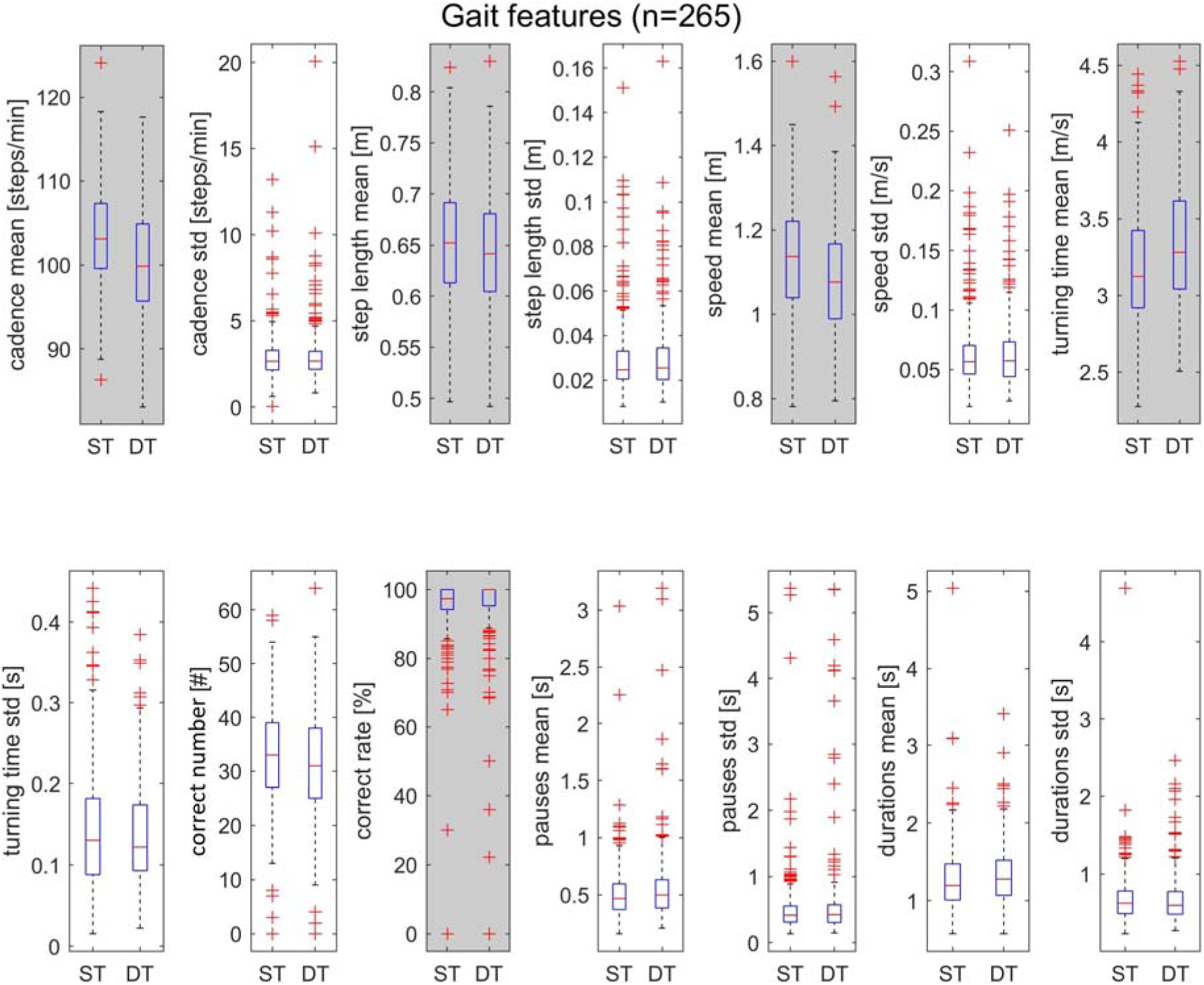
Single-task (ST) and dual-task (DT) features distribution for the gait test. Grey areas indicates features that are significantly different between ST and DT (*p* < 0.05).

For the balance features, significant differences between ST and DT performance were found for all the features (p < 0.05) except the estimated *root mean square* in the AP direction (*p*= 0.11) and the s*pectral centroid* in the AP direction (*p* = 0.76). The cognitive features were not significantly different between ST and DT for balance tests (all *p* > 0.06), except for the *durations std (p* = 0.004) (Figure 2).

**Figure 2:**
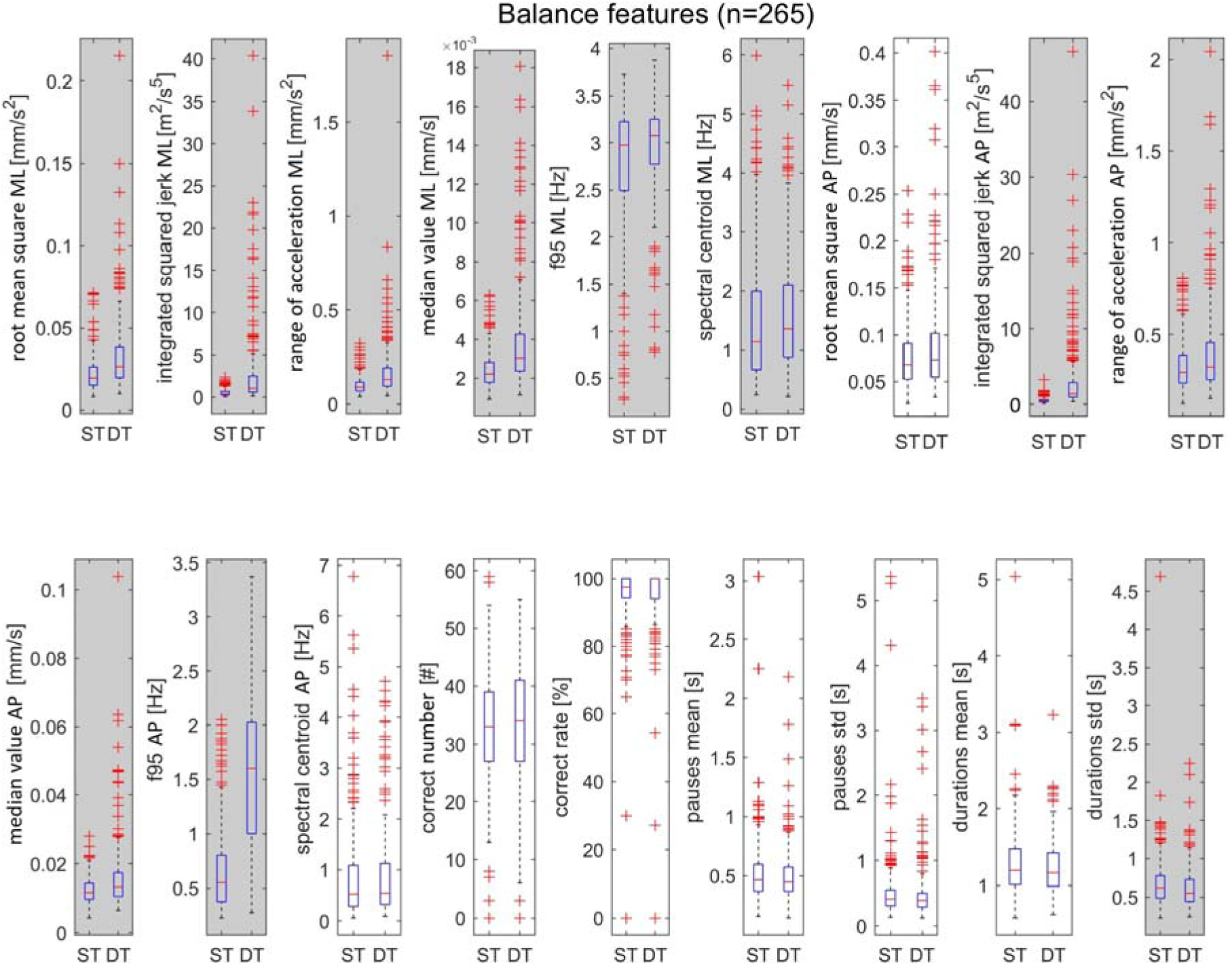
Single-task (ST) and dual-task (DT) features distribution for the balance test. Balance features are calculated for medio-lateral (ML) and antero-posterior (AP) directions. Grey areas indicates features that are significantly different between ST and DT (*p* < 0.05).

### 3.2 Intra-subject variability

Twenty-three players performed two baseline tests in the first month of the season. The paired Wilkoxon signed rank tests showed no significant DT effect differences for the gait and the balance, except for the *step length std* (*p* = 0.030, d = 0.37).

Twenty-eight players who did not sustain a concussion performed a baseline before the start of the season and a baseline at the end of the season (i.e. more than 18 weeks after the beginning of the season. The paired Wilkoxon signed rank tests showed a significantly increased DT effect for *correct number* during balance (*p* = 0.025, *d* = 0.42) and gait (*p* < 0.001 *d* = 0.38), the *pauses std* during gait (*p*= 0.022, *d* = 0.45), the *durations std* during gait (*p*= 0.021, *d* = 0.47), and the *correct rate* during gait (*p* = 0.002, *d* = 0.19).

### 3.3 Intra-subject concussion analysis

Thirteen players who made a baseline before the start of the season sustained a concussion during the season. The paired Wilkoxon signed rank tests showed no significant DT effect differences during balance (Table 2). For the DT effect differences during gait, the effect of *speed std* (*p* = 0.036, *d* = 0.62), the effect of *correct answe*r (*p* = 0.021, *d* = 0.75), the effect of *pauses mean* (*p* = 0.031, *d* = 0.69), and the effect of *durations std* (*p* = 0.040 *d* = 0.71) showed significant differences (Figure 3). The calculated total DT effect showed a significant increase after the concussions compared to their respective baselines (*p* < 0.001, *d* = 1.5). Nine of the 13 players performed a DT test between five and seven days after the concussion. Their total DT effect was still significantly higher than their baseline (*p* = 0.038, *d* = 0.65). Figure 4A indicates the probability of sustaining a concussion based on the DT test outcome for a player that performed a pre-season baseline.

**Figure 3:**
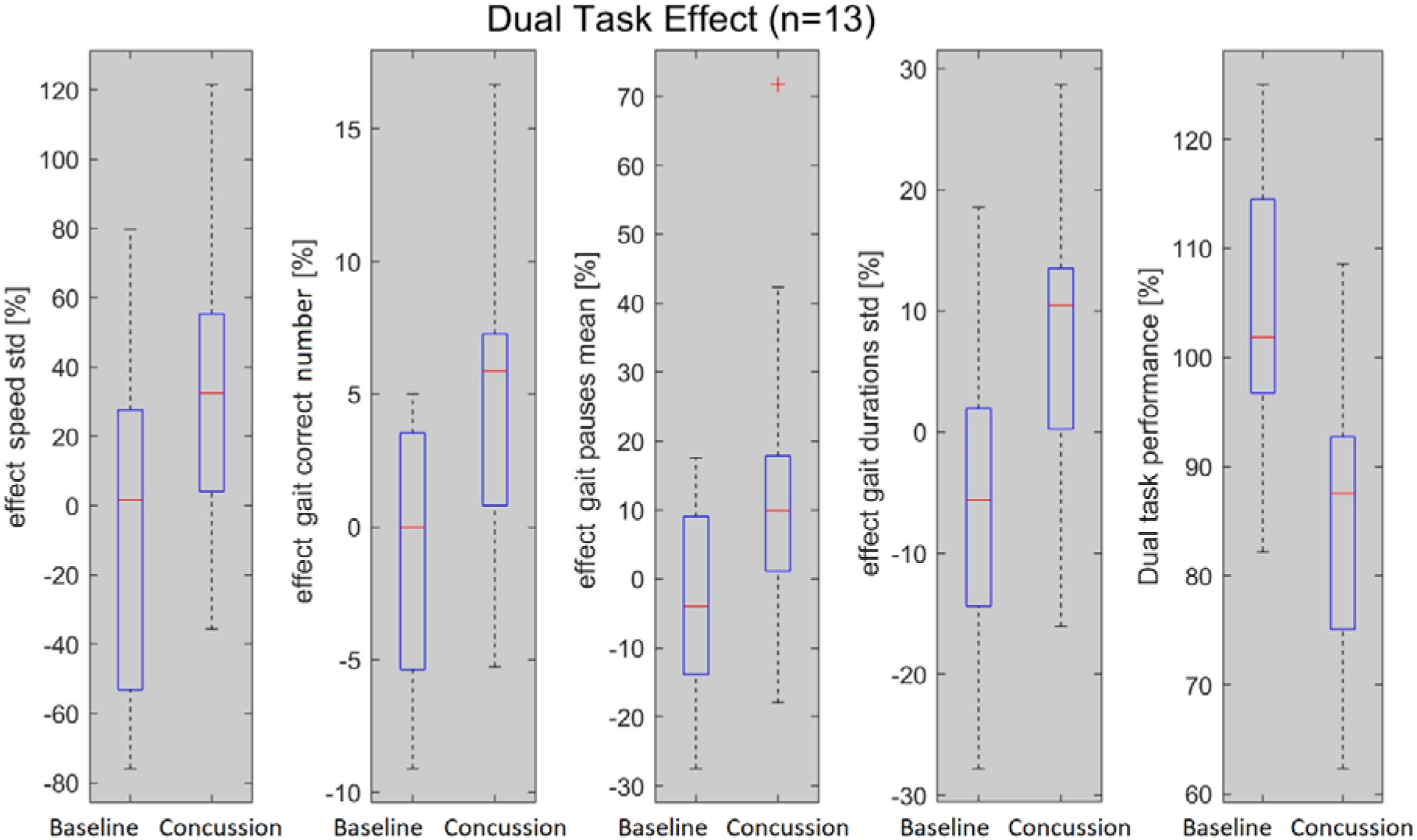
Dual-task effects for the features significantly differed between the baselines performed before the start of the season and their corresponding concussion sustained during the season. The Dual-task performance is 100 minus the mean of the four significant feature effects.

**Figure 4:**
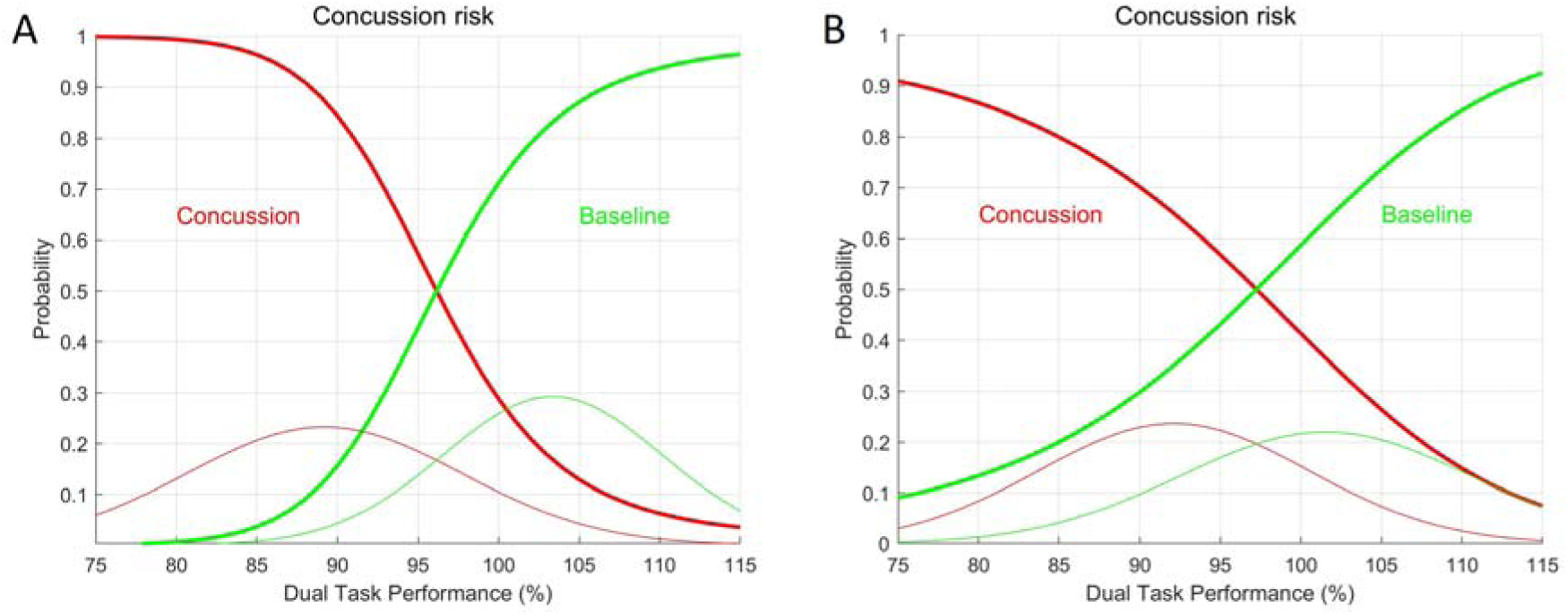
Probability of sustaining a concussion based on the calculated total Dual Task effect. **A.** A 5% decrease in the Dual Task test performance compared to the athlete pre-season baseline give a probability of 70% to have a concussion. A 10% decrease in the Dual Task performance effect give a concussion risk of more than 95%. **B.** A 5% decrease in the Dual Task performance compared to the overall baselines give a probability of 65% to have a concussion. A 10% decrease in the Dual Task performance give a concussion risk of 80%.

**Table 2:** Comparison of the dual task effect for the 13 hockey players who sustained a concussion during the season and their pre-season baseline.

| Dual task effect (%) | Baseline<br>mean ± sd | Concussion<br>mean ± sd | *p* |
| --- | --- | --- | --- |
| **Balance features** |  |  |  |
| root mean square ML | 8,50 ± 27,82 | 17,08 ± 21,02 | 0,505 |
| integrated squared jerk ML | 30,94 ± 32,52 | 36,10 ± 26,47 | 0,608 |
| range of acceleration ML | 11,72 ± 26,33 | 19,58 ± 19,61 | 0,412 |
| median value ML | 18,17 ± 21,67 | 19,18 ± 19,54 | 0,644 |
| F95 ML | 3,04 ± 21,65 | 6,90 ± 17,35 | 0,608 |
| spectral centroid ML | -2,90 ± 36,56 | 14,83 ± 30,19 | 0,238 |
| root mean square AP | 2,68 ± 18,54 | 6,51 ± 30,10 | 0,837 |
| integrated squared jerk AP | 48,53 ± 18,24 | 48,28 ± 19,39 | 1,000 |
| range of acceleration AP | 4,54 ± 15,37 | 8,63 ± 28,17 | 0,682 |
| median value AP | 14,20 ± 16,87 | 13,27 ± 13,81 | 0,608 |
| F95 AP | 29,40 ± 37,05 | 23,50 ± 37,81 | 0,626 |
| spectral centroid AP | 10,64 ± 39,02 | 25,46 ± 37,83 | 0,330 |
| balance correct number | 3,72 ± 7,76 | 0,23 ± 4,60 | 0,069 |
| balance correct rate | 1,41 ± 3,66 | 1,07 ± 2,04 | 0,749 |
| balance pauses mean | -5,79 ± 16,75 | 6,70 ± 15,81 | 0,081 |
| balance pauses std | 0,94 ± 23,66 | 4,61 ± 24,10 | 0,538 |
| balance durations mean | -2,16 ± 6,21 | -0,60 ± 8,00 | 0,758 |
| balance durations std | -6,52 ± 19,78 | -1,55 ± 13,17 | 0,473 |
| **Gait features** |  |  |  |
| cadence mean | -1,53 ± 1,80 | -1,54 ± 2,10 | 1,000 |
| cadence std | 6,93 ± 32,78 | 9,96 ± 20,67 | 0,356 |
| step length mean | -1,27 ± 2,06 | -1,47 ± 2,57 | 0,758 |
| step length std | -1,79 ± 27,94 | 19,26 ± 27,75 | 0,112 |
| speed mean | -2,81 ± 3,57 | -2,97 ± 4,19 | 0,798 |
| speed std | -3,61 ± 24,17 | 17,56 ± 23,50 | 0,036 |
| turning time mean | 2,37 ± 1,93 | 2,39 ± 2,01 | 0,959 |
| turning time std | -9,98 ± 23,52 | -2,36 ± 14,27 | 0,473 |
| gait correct number | -0,75 ± 5,00 | 4,69 ± 6,16 | 0,021 |
| gait correct rate | 1,11 ± 3,66 | -0,41 ± 4,24 | 0,300 |
| gait pauses mean | -4,00 ± 14,27 | 13,94 ± 22,56 | 0,031 |
| gait pauses std | -0,59 ± 19,88 | 16,96 ± 28,11 | 0,112 |
| gait durations mean | 2,23 ± 6,87 | 2,64 ± 7,64 | 0,837 |
| gait durations std | -5,05 ± 13,14 | 7,08 ± 12,80 | 0,040 |
| total dual task effect | -0,75 ± 5,00 | 4,69 ± 6,16 | < 0,001 |

DCom is the displacement of the center of mass ML is medio-lateral direction and AP is antero-posterior direction. Grey areas indicates features that are significantly different between Baseline and Concussion tests ( $p < 0.05$ ).

### 3.4 Inter-subject concussion analysis

Sixty-three players made a baseline test before the start of the season but did not get a concussion. This group was compared with the group of 15 players who sustained a concussion during the season. The paired Wilkoxon signed rank tests significantly increased the total DT effect (*p* = 0.001, *d* = 1.4). Figure 4B indicates the probability of sustaining a concussion based on the DT test outcome for players who did not perform a baseline.

## 4. Discussion

The objective of this study was to assess the capacity of a DT test to identify hockey players who sustained a concussion. First, most of the features showed a higher effect for DT than for ST (figure 1 and 2), supporting results of previous studies highlighting the need for shared brain attention when performing two different tasks ^15,16^. The DT affected the gait and the balance features, but not the cognitive features. This can be explained through a cognition-first strategy adopted intuitively by the players during the baselines, as the experimenter provided no specific indications concerning prioritization. A similar observation was highlighted by Yogev-Seligmann et al. ^39^.

For the 23 players that performed two baseline assessments before the season, only the *step length std* showed a significant difference between the two tests but with a weak effect size. It indicates that there is no learning effect of the procedure by the hockey players. A similar outcome was highlighted for a healthy control group when analyzing the reliability of spatio-temporal parameters in people with multiple sclerosis ^40^. In our study, no correction for multiple variables testing was applied to the *p*-values to provide a more conservative outcome. In this context, obtaining only one feature with a *p-*value lower than 0.05 indicates a good test-retest consistency.

The comparison of the 28 un-concussed players that performed a DT test before the start of the season and at the end of the season indicates that the effects of five features (i.e. the *correct number* during balance and during gait, the *correct rate*, the *pauses std* and *duration*s *std* during gait) increased at the end of the season, with a very week or weak effect size. This outcome may indicate slight mental fatigue of the athletes at the end of the season, as observed for cognitive performance under DT conditions after an acute fatigue protocol ^41^.

The tests performed right after a concussion compared to their corresponding baseline highlighted significant increases in the DT effects with moderate effect size for the gait *speed std*, the *correct number*, the *pause mean* and the *durations std* during gait. It resulted in a significant increase in total DT effect for the concussion group, with a very strong effect size. These results provide clear evidence that the DT test applied in this study can identify the acute effect of concussion on DT performance compared to its own baseline. The effect was still observed one week after the concussion. Similar results were observed after 8,5 days in a study where participants had to talk during walking ^42^. In our study, the cognitive features were the most affected by the concussion during gait, showing that after a concussion, hockey players are not able to keep their cognitive level while walking. This outcome also aligns with the gait-first strategy observed in healthy adults ^20^. Moreover, the gait *speed std* was also affected, meaning that both tasks (gait and cognitive) were saturated when performing the DT with a concussion. The fact that no effect differences were observed during the static balance task could indicate that this part needed to be more challenging to observe changes with elite players. For example, it was shown that older adults have lower dual-task performance than younger adults ^16^. The difficulty of the static balance could be further enhanced by performing a tandem stance or with closed eyes to see if increasing the difficulty of the motor task affects the outcome of the DT after a concussion. A recent review highlighted that the complexity of the cognitive task was not affecting healthy individuals, but differences were found depending on the age of the participants and the kind of postural task ^43^. Moreover, other studies found significant DT effects for balance and cognitive tasks with concussed patients ^14,44^.

The model built using the DT performance to estimate concussion risk based on the normal distribution of the baseline and the concussion groups provides a practical tool for in-field use. Indeed, the outcome of a DT test performed by an athlete can be directly interpreted using the paired model (i.e., if the player has a baseline) or the global model. As observed in Figure 4A, a decrease of 5% in the DT performance of a test compared to its baseline gives a detection rate of 70% of the concussions. When the DT performance decrease by 10%, the detection rate reaches 95. If the athlete don’t have a baseline to compare with, a 5% decrease in the DT performance gives a detection rate of to 65%. Therefore, the paired model (i.e. when the athlete has a baseline) is more precise and should be promoted for regular use. The model also indicates the risk of missing a concussion, as the normal distribution of the baselines and the concussion intersect. With the paired model, there is a risk of 30% missing a concussion if the DT performance is identical between the two tests. Increasing the number of DT tests included in the model with new data could progressively improve the robustness of the model, as the normal distribution of the two groups will be more precise.

Improving the global model would also be very useful for in-field application, as performing baselines regularly with athletes is sometimes challenging. Moreover, working with professional athletes can be difficult, as they have tight schedules, and everything has to serve a quantifiable purpose. Conducting these DT tests in a non-clinical environment could have increased variability in results, underscoring the need for both rigor and adaptability in test administration. However, this study demonstrates feasibility and validity of the test, which is very important, as in-field analysis is essential for coaches to better understand real-world dynamics ^45^. We also emphasize the practicality and usefulness of the test by providing a performance index for the DT centred on 100% that is easily understandable by practitioners, as this is key for proper use in the field ^46^.

Finally, higher sensitivity may be reached by looking at the concussion history of the hockey players, as this can affect DT performance ^47–49^. Long-term effects of a concussion are also essential to consider and monitor, as too early return to play can lead to disastrous consequences for the athlete^50,51^. As observed in the current study, the DT test proposed in this study could help better understand these long-term effects for even longer than one week. It has been shown that DT tests can detect remaining cognitive, balance and gait disorders for several weeks after a concussion, whereas ST tests only allow for the detection of acute effects ^48,52,53^. Further investigations should implement DT follow-up tests for up to one month after the concussion.

## 5. Conclusion

Dual-task tests are effective in detecting and monitoring concussions in elite ice hockey players. When performing a DT test right after a concussion, athletes showed reduced cognitive performance while walking and this impairment remained noticeable even one week later. Finally, individual and global models can estimate concussion risk based on DT test outcomes, enabling efficient on-field screening of athletes.

## Data Availability

All data produced in the present study are available upon reasonable request to the authors

## Acknowledgement

The authors want to thanks all the players that participated to this study. This project was funded by the Innosuisse (grant number 100.098 IP-ICT).

## Authors’ contributions

FM designed the study, processed the data, performed the statistical analysis and drafted the manuscript; NB carried out the tests and processed the data; DS carried out the tests and helped to draft the manuscript: LC participated to the data processing and analysis; MF conceived and coordinated the study and helped draft the manuscript. AI conceived the study, and participated in its design and helped to draft the manuscript. All authors have read and approved the final version of the manuscript, and agree with the order of presentation of the authors.

